# Normative cognitive scores in western India, stratified by age, rurality, cognitive domains, and psychiatric comorbidity

**DOI:** 10.1101/2022.03.22.22272678

**Authors:** Uma Sundar, Amita Mukhopadhyay, Suresh Sundar, Nilesh Shah

**Author notes:** **Corresponding Author:** Dr Amita Mukhopadhyay.

## Abstract

**Background and objectives:** Indian ethnic and educational diversities necessitate obtaining normative cognitive data in different populations. We aimed to evaluate cognitive scores using a Marathi translation of the Kolkata Cognitive Battery (KCB), and to study the association of KCB components with depression and socio-demographic variables.

**Methodology:** We studied 2651 individuals aged ≥40 years, without pre-existing neuropsychiatric conditions, from urban (Mumbai) and rural districts of Maharashtra. For each component of KCB, the lowest 10th percentile score was used to define cognitive impairment.

**Results:** We studied 1435 (54%) rural and 1216 (46%) urban residents equally divided by gender (1316 women, 1335 men), average age 54 years. KCB scores were significantly lower with female sex, older age, illiteracy and depression. The largest effect sizes attributable to these factors were in the domains of calculation (gender), visuo-constructional ability (VCA; rurality) and verbal fluency (VF; depression). Scores remained significantly lower in rural residents after controlling for age, sex and education, particularly for VCA, immediate recall and calculation.

**Conclusion:** This Marathi KCB having been validated on large urban as well as rural samples, may be used to study cognition in Marathi speaking populations with appropriate cut-offs tailored to the degree of urbanization of the population.

## Background

Community based studies on cognition in India, especially in rural areas, are few. Ethnic, socio-cultural, educational and linguistic diversities in India make it necessary to obtain normative data in cognition in different populations. Tripathi et al have reported objective cognitive dysfunction in the areas of “orientation” and “concentration” and “functioning/self-care” in normally aging older adults in urban Lucknow.^1^ Gambhir et al, in a rural study of elderly, found a Dementia prevalence of 2.7% and suggested that Hindi Mental State Examination (HMSE) score of 17 should be used as cutoff for the illiterate population.^2^ Ganguli et al described a battery of cognitive tests that was developed for screening an illiterate elderly rural Hindi-speaking population, based on their research conducted in Ballabhgarh district in Haryana.^3^ Further, Banerjee et al^4^ and Das et al^5^ have obtained normative data for cognitive functions in a Bengali urban population utilizing the Kolkata Cognitive Screening Battery (KCB).

The mini mental state examination (MMSE) and the Hindi mental state examination (HMSE), although popular and easy to administer, suffer from a lack of sensitivity to early dementia, especially of the subcortical variety.^6^ We aimed to evaluate cognitive scores using the KCB, in an age-stratified manner, in Maharashtrian rural and urban communities, and evolve cut-off scores based on individual test scores within the KCB, as a step towards refining its sensitivity and specificity as a screening instrument to detect cognitive impairment in the specific population under study. Further, we aimed to compare differences in individual test scores classed according to different socio-demographic variables as well as Geriatric Depression Scale (GDS)^7^ scores and finally, compare our sample’s scores with those of the Kolkata and Ballabhgarh study populations.

## Methodology

### Study design

The KCB has been validated in urban populations by Das et al,^5^ and the Hindi cognitive battery from which it was derived, has been validated in the rural setting by Ganguli et al.^3^ In the first part of this study, the partially modified Hindi cognitive screening battery used by Ganguli et al was translated into Marathi and again back - translated to Hindi by two independent bilingual translators. The GDS was also translated into Marathi and back-translated into Hindi. Field testing on 40 persons was done initially, for both KCB and GDS, to ensure inter and intra investigator reliability, being scored by the chief investigator and a qualified psychologist each time. Intra-rater reliability was 1, and inter-rater reliability was over 0.9 for both KCB and GDS. Qualified psychologists and final year residents in psychiatry training were trained in administration of the translated KCB and GDS test by the chief investigator and a consultant psychiatrist.

In the second part, a cross sectional interview-based survey was conducted in urban, semi urban and rural areas of Maharashtra.

### Setting

The study was conducted in the villages of Vaitarna (Vadataluka, Thane district, population 671 as per 2011 census), Birwadi (Shahapurtaluka, Thane district, population 3415), and Penn taluka (Raigad district, population 37,852) over a period of 16 months from March 2015 to July 2016. These areas were chosen considering that they are the setting of ongoing primary health care projects under the community health department of the parent hospital, which made it feasible to obtain information on household numbers, size and the number of people over 40 years of age in these areas. The urban areas in Mumbai included Kumbharwada in Dharavi, Kala Chowki and Antop Hill areas.

### Data collection

All persons aged 40 and over and residing in the study areas were invited to take part in the survey. We excluded persons with known mental retardation, previous stroke, any known psychiatric illness, or those reported by family to have previously diagnosed dementia or significant memory impairment impeding daily activities. Healthy individuals, who met these criteria and consented to take part in the study, comprised the final sample.

Trained personnel first recorded all epidemiological and demographic data for the consenting respondents, and then administered the KCB. We used the KCB version validated by Ganguli et al, as it had questions on post office, district, village and block, relevant to a rural community.^3^

The Marathi KCB was validated by re-administering 30% of the cognition tasks from each of the eight domains of the KCB on 40 randomly chosen study participants—by a second field researcher for inter-rater, and by the same researcher after a gap of 6 weeks for intra-rater reliability. Test administration and scoring were supervised by the chief investigator.

Our primary outcome measure was the age-stratified KCB score. Data collected on completed years of schooling was classified into educational categories namely, illiterate, primary (1-5 years), secondary (6-12 years), and graduation level (13 years and above).

#### Cut-off scores

For screening a large population, operational cutoff points are essential to differentiate between the cognitively impaired and the unimpaired groups.^8^ Following the precedent set by Das et al,^5^ we chose the lowest 10th percentile score as the cut-off point to identify the significantly cognitively impaired tenth of the population. In GDS, scores range from 0-30. The higher scores indicate undetected depression. We chose the 90th percentile as the operational cut-off point to identify respondents with masked depression.

### Statistical methods

The data was analyzed using appropriate SPSS^9^ and JASP.^10^ Values were expressed in frequencies as well as percentages in relevant rows. Spearman rank correlation coefficients were calculated to determine intra and inter observer reliability. Anova was used to compare means between groups. Kruskal Wallis H test was used to analyze overall differences between age groups and educational classes, and Mann Whitney U test was used to compare the two genders, urban and rural groups, groups with and without masked depression, and to compare each of the subgroups in age and education with the other subgroups.

The study was cleared by the Institutional Ethical Committee of Lokmanya Tilak Municipal Medical College and General Hospital, Sion, Mumbai.

## Results

### Demographic profile

We screened 2752 persons over 40 years of age; we excluded 19 persons with known intellectual developmental disorder, 31 with possible previous stroke, 36 with previous psychiatric illness, and 15 persons reported by family to have previously diagnosed dementia or significant memory impairment impeding daily activities. Our final sample size was 2651 individuals who satisfied the inclusion and exclusion criteria and consented to participate in an interview. Their socio-demographic characteristics are summarized in Table 1. Among these participants, 1435 (54%) were from rural localities while 1216 (46%) belonged to urban localities. The study group was equally divided with respect to gender, comprising 1335 men and 1316 women. However, in the younger age groups (<60 years), men outnumbered women and this trend was reversed for the older age groups. The mean age of the study participants was 54 years and women were slightly older (mean 54.28 years) than men (mean 53.87 years), though this difference was not significant. Most of the participants were poorly educated. Out of 1335 men, 752 (56.3%) had studied for five years or less, and among women the corresponding proportion was significantly higher (1001 of 1316, 76.1%, p<0.01). Correspondingly, men significantly outnumbered women in the groups with higher education. We also noted a significant gender difference in occupation, with most of the women participants reporting as housewives (1108, 84.2% of 1316) and most of the remainder in unskilled and semiskilled occupations. Among men, 516/1335, i.e., 38.4%, reported employment in unskilled vocations, followed by semiskilled and clerical work respectively. We observed a significant downtrend in mean years of education with increasing age (table 1). We observed a significant difference in scores between rural and urban subsets of the study sample. The urban residents were significantly younger and had significantly higher years of education than the rural residents (table 1). While the study sample comprised roughly equal numbers of men and women with no significant difference in their mean age, the two gender groups differed notably in education, with men on average having 6.5 years of schooling compared to 4.7 years for women (table 1).

**Table 1:**
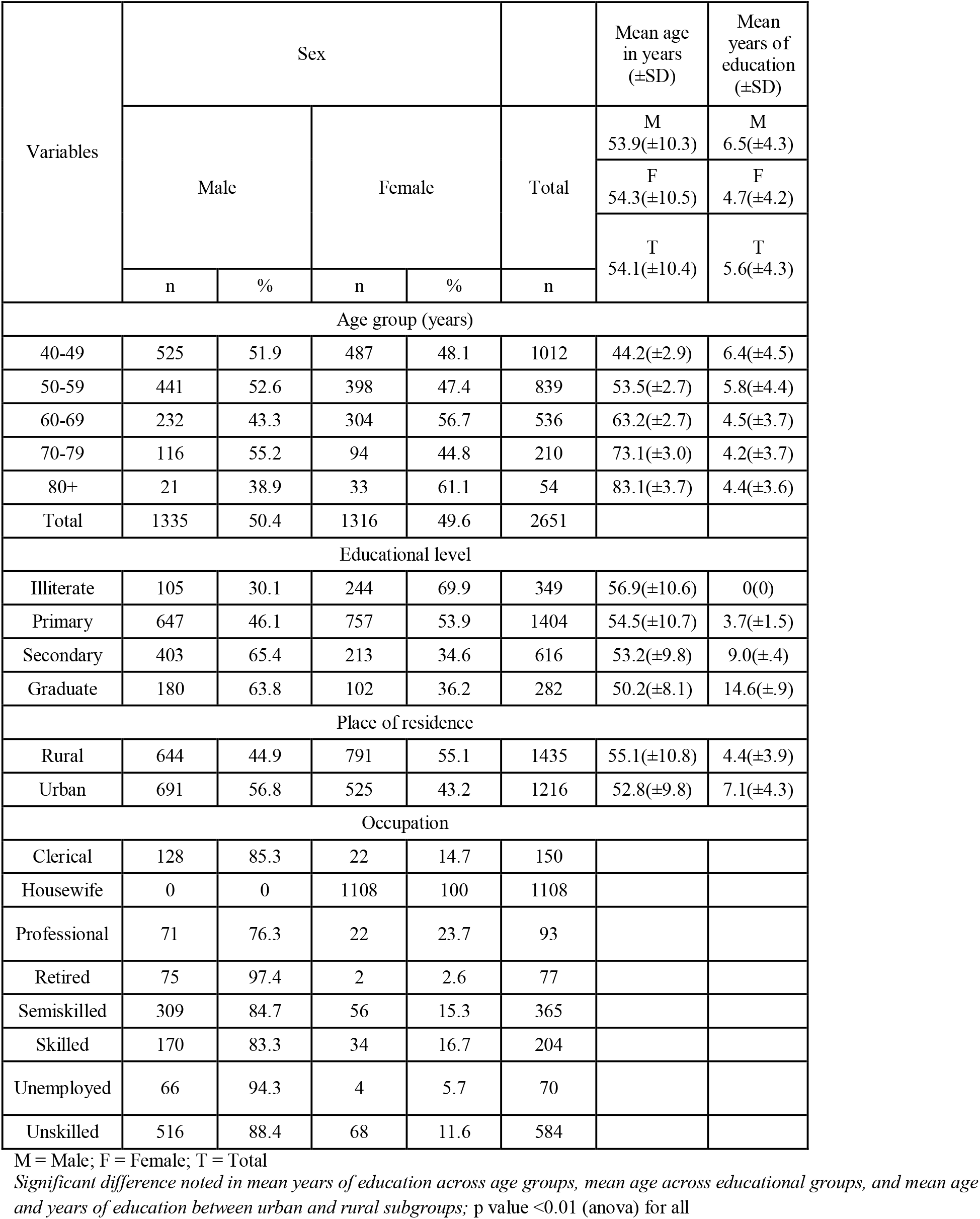
Demographic characteristics

### Test scores and validation

Individual test scores with mean, median, standard deviation, interquartile range and 10th and 90th percentile scores are recorded in table 2. Spearman rank correlation coefficients for both inter and intra rater reliability were high for each test, ranging from 0.92 to 1.0. The score range for each domain spanned from zero to the maximum attainable score, except in the case of immediate recall, in which the maximum score attained was 28 out of 30.

**Table 2:** Overview of test scores

| KCB<br>Component<br>Tests | Maximum<br>score | n | Range | Mean | Std.<br>Deviation | Median | Inter-<br>quartile<br>range | 10<sup>th</sup><br>percentile<br>score | 90<sup>th</sup><br>percentile<br>score |
| --- | --- | --- | --- | --- | --- | --- | --- | --- | --- |
| Test 1 Verbal<br>Fluency | NS | 2651 | 0-32 | 11.86 | 5.35 | 12 | 7 | 5 | 20 |
| Test 2 Object<br>Naming | 15 | 2650 | 0-15 | 10.00 | 3.80 | 10 | 8 | 5 | 15 |
| Test 3 Mental<br>State<br>Examination | 30 | 2635 | 0-30 | 24.56 | 5.31 | 26 | 8 | 17 | 30 |
| Test 4<br>Calculation | 5 | 2567 | 0-5 | 3.45 | 1.69 | 4 | 3 | 1 | 5 |
| Test 5<br>Immediate<br>Recall | 30 | 2651 | 0-28 | 12.01 | 5.89 | 12 | 8 | 4 | 20 |
| Test 6 Visuo-constructional<br>Ability | 13 | 2646 | 0-13 | 6.68 | 3.79 | 6 | 6 | 2 | 12 |
| Test 7<br>Delayed<br>Word List | 10 | 2623 | 0-10 | 4.77 | 2.14 | 5 | 2 | 2 | 7 |
| Test 8<br>Delayed<br>Recognition | 20 | 2637 | 0-20 | 8.57 | 4.36 | 9 | 6 | 2 | 14 |

### Gender

The individual test scores refined by gender, shown in table 3, highlight a significant difference in the mean scores, with women scoring lower than men on each test. The 90^th^ percentile scores were higher for men in tests of verbal fluency, immediate recall, visuo-constructional ability and delayed recognition, and the 10^th^ percentile scores were higher for MMSE, visuo-constructional ability and delayed recognition. The difference in test scores attributable to gender, i.e., effect size, ranged from 5% to 20%, with the largest effect size observed for test 4, calculation.

**Table 3:** Test scores sorted by gender and their comparison by Mann-Whitney U test

| Parameters | Gender | n | Mean | Std. Deviation | 10<sup>th</sup> percentile score | 90<sup>th</sup> percentile score | p value | Effect size* | 95% CI for effect size |  |
| --- | --- | --- | --- | --- | --- | --- | --- | --- | --- | --- |
|  |  |  |  |  |  |  |  |  | Lower | Upper |
| Test 1 Verbal Fluency | Men | 1335 | 12.36 | 5.4 | 5 | 20 | <0.01 | 0.113 | 0.070 | 0.157 |
|  | Women | 1316 | 11.35 | 5.25 | 5 | 19 |  |  |  |  |
| Test 2 Object Naming | Men | 1335 | 10.16 | 3.62 | 5 | 15 | <0.05 | 0.051 | 0.007 | 0.095 |
|  | Women | 1315 | 9.85 | 3.97 | 5 | 15 |  |  |  |  |
| Test 3 Mental State Examination | Men | 1324 | 25.17 | 5.21 | 17 | 30 | <0.01 | 0.16 | 0.117 | 0.203 |
|  | Women | 1311 | 23.94 | 5.35 | 16 | 30 |  |  |  |  |
| Test 4 Calculation | Men | 1282 | 3.74 | 1.6 | 1 | 5 | <0.01 | 0.203 | 0.16 | 0.246 |
|  | Women | 1285 | 3.15 | 1.72 | 1 | 5 |  |  |  |  |
| Test 5 Immediate Recall | Men | 1335 | 12.75 | 5.92 | 4 | 20 | <0.01 | 0.144 | 0.101 | 0.187 |
|  | Women | 1316 | 11.27 | 5.76 | 4 | 19 |  |  |  |  |
| Test 6 Visuo-constructional Ability | Men | 1332 | 7.13 | 3.76 | 2 | 12 | <0.01 | 0.139 | 0.095 | 0.181 |
|  | Women | 1314 | 6.23 | 3.78 | 0 | 11 |  |  |  |  |
| Test 7 Delayed Word List | Men | 1319 | 4.93 | 2.07 | 2 | 7 | <0.01 | 0.089 | 0.045 | 0.132 |
|  | Women | 1304 | 4.61 | 2.19 | 2 | 7 |  |  |  |  |
| Test 8 Delayed Recognition | Men | 1327 | 8.91 | 4.32 | 3 | 14 | <0.01 | 0.085 | 0.041 | 0.129 |
|  | Women | 1310 | 8.22 | 4.35 | 2 | 13 |  |  |  |  |

\* For the Mann-Whitney test, effect size is given by the rank biserial correlation

### Rural vs Urban residence

The individual test scores for rural and urban subsets, shown in table 4, demonstrate higher mean scores in each test for the urban subset. Tenth percentile scores were higher for urban residents in all tests except delayed word list, and 90^th^ percentile scores were also higher in four of the eight tests. Effect size ranged from 10% to 47%, with the largest difference observed for test 6, visuo-constructional ability followed by test 5, immediate recall.

**Table 4:** Test scores sorted by urban/rural residence, and their comparison by Mann-Whitney U test

| Parameters | Residence | n | Mean | Std. Deviation | 10<sup>th</sup> percentile score | 90<sup>th</sup> percentile score | p value | Effect size* | 95% CI for effect size |  |
| --- | --- | --- | --- | --- | --- | --- | --- | --- | --- | --- |
|  |  |  |  |  |  |  |  |  | Lower | Upper |
| Test 1 Verbal Fluency | Rural | 1435 | 10.55 | 5.27 | 5 | 18 | <0.01 | -0.326 | -0.365 | -0.286 |
|  | Urban | 1216 | 13.41 | 5.02 | 7 | 20 |  |  |  |  |
| Test 2 Object Naming | Rural | 1434 | 9.68 | 4.25 | 4 | 15 | <0.01 | -0.109 | -0.153 | -0.065 |
|  | Urban | 1216 | 10.39 | 3.15 | 6 | 15 |  |  |  |  |
| Test 3 Mental State Examination | Rural | 1427 | 23.14 | 5.33 | 15 | 30 | <0.01 | -0.381 | -0.419 | -0.343 |
|  | Urban | 1208 | 26.23 | 4.78 | 19 | 30 |  |  |  |  |
| Test 4 Calculation | Rural | 1424 | 2.94 | 1.73 | 0 | 5 | <0.01 | -0.378 | -0.416 | -0.339 |
|  | Urban | 1143 | 4.07 | 1.41 | 2 | 5 |  |  |  |  |
| Test 5 Immediate Recall | Rural | 1435 | 9.87 | 5.18 | 4 | 16 | <0.01 | -0.466 | -0.5 | -0.43 |
|  | Urban | 1216 | 14.54 | 5.66 | 7 | 22 |  |  |  |  |
| Test 6 Visuo-constructional Ability | Rural | 1432 | 5.27 | 3.43 | 0 | 10 | <0.01 | -0.472 | -0.505 | -0.437 |
|  | Urban | 1214 | 8.34 | 3.52 | 3 | 12 |  |  |  |  |
| Test 7 Delayed Word List | Rural | 1427 | 4.5 | 2.17 | 2 | 7 | <0.01 | -0.168 | -0.211 | -0.125 |
|  | Urban | 1196 | 5.09 | 2.06 | 2 | 7 |  |  |  |  |
| Test 8 Delayed Recognition | Rural | 1435 | 7.78 | 4.46 | 2 | 13 | <0.01 | -0.248 | -0.289 | -0.206 |
|  | Urban | 1202 | 9.51 | 4.06 | 4 | 14 |  |  |  |  |

\* For the Mann-Whitney test, effect size is given by the rank biserial correlation

### Presence vs Absence of depression

In our study sample, the 90th percentile score for GDS was 17, which we used as the operational cut off to identify masked depression. We observed significantly lower mean scores on all tests among the respondents who scored 17 and above on the GDS (table 5). The largest effect size, up to 18%, was seen in Test 3, Mental State Examination, and Test 1, Verbal Fluency.

**Table 5:** Test scores sorted by presence/absence of masked depression, and their comparison by Mann-Whitney U test

| Parameters | Depression<sup>#</sup> | n | Mean | Std. Deviation | 10<sup>th</sup> percentile score | 90<sup>th</sup> percentile score | p value | Effect size* | 95% CI for effect size |  |
| --- | --- | --- | --- | --- | --- | --- | --- | --- | --- | --- |
|  |  |  |  |  |  |  |  |  | Lower | Upper |
| Test 1 Verbal Fluency | No | 2376 | 12.03 | 5.41 | 5 | 20 | <0.01 | 0.173 | 0.102 | 0.242 |
|  | Yes | 275 | 10.39 | 4.49 | 5 | 16 |  |  |  |  |
| Test 2 Object Naming | No | 2375 | 10.11 | 3.76 | 5 | 15 | <0.01 | 0.141 | 0.07 | 0.211 |
|  | Yes | 275 | 9.09 | 4.01 | 5 | 15 |  |  |  |  |
| Test 3 Mental State Examination | No | 2364 | 24.71 | 5.27 | 17 | 30 | <0.01 | 0.187 | 0.116 | 0.256 |
|  | Yes | 271 | 23.25 | 5.54 | 16 | 29 |  |  |  |  |
| Test 4 Calculation | No | 2299 | 3.49 | 1.68 | 1 | 5 | <0.01 | 0.152 | 0.08 | 0.222 |
|  | Yes | 268 | 3.06 | 1.76 | 0 | 5 |  |  |  |  |
| Test 5 Immediate Recall | No | 2376 | 12.20 | 5.89 | 5 | 20 | <0.01 | 0.165 | 0.094 | 0.234 |
|  | Yes | 275 | 10.40 | 5.60 | 3 | 18 |  |  |  |  |
| Test 6 Visuo-constructional Ability | No | 2371 | 6.78 | 3.76 | 2 | 12 | <0.01 | 0.135 | 0.063 | 0.205 |
|  | Yes | 275 | 5.81 | 4.01 | 0 | 11 |  |  |  |  |
| Test 7 Delayed Word List | No | 2351 | 4.84 | 2.12 | 2 | 7 | <0.01 | 0.159 | 0.087 | 0.228 |
|  | Yes | 272 | 4.21 | 2.25 | 1 | 7 |  |  |  |  |
| Test 8 Delayed Recognition | No | 2362 | 8.68 | 4.33 | 3 | 14 | <0.01 | 0.136 | 0.065 | 0.206 |
|  | Yes | 275 | 7.59 | 4.58 | 2 | 12 |  |  |  |  |

\* For the Mann-Whitney test, effect size is given by the rank biserial correlation
<sup>#</sup>Presence of depression operationally defined as GDS score $\geq 17$

Table 6 shows the comparison of mean scores between different educational groups. Significant differences were observed in all tests between the illiterate group and every other educational category, except for the comparison between illiterate and primary education group for object naming and delayed recognition tests. Similarly, in the comparison between secondary education and graduate level groups, the difference was not significant for several tests including immediate recall, visuo-constructional ability and delayed recognition.

**Table 6:** Test scores of each educational level compared with higher and lower levels using Mann Whitney U test

| Parameters | Illiterate vs<br>Primary | Illiterate vs<br>Secondary | Illiterate vs<br>Graduate | Primary vs<br>Secondary | Primary vs<br>Graduate | Secondary<br>vs Graduate |
| --- | --- | --- | --- | --- | --- | --- |
| Test 1 Verbal Fluency | P<0.01 | P<0.01 | P<0.01 | P<0.01 | P<0.01 | P<0.05 |
| Test 2 Object Naming | NS | P<0.01 | P<0.01 | P<0.01 | P<0.01 | P<0.01 |
| Test 3 Mental State<br>Examination | P<0.01 | P<0.01 | P<0.01 | P<0.01 | P<0.01 | P<0.01 |
| Test 4 Calculation | P<0.01 | P<0.01 | P<0.01 | P<0.01 | P<0.01 | P<0.01 |
| Test 5 Immediate Recall | P<0.01 | P<0.01 | P<0.01 | P<0.01 | P<0.01 | NS |
| Test 6 Visuo-constructional Ability | P<0.01 | P<0.01 | P<0.01 | P<0.01 | P<0.01 | NS |
| Test 7 Delayed Word List | P<0.01 | P<0.01 | P<0.01 | NS | P<0.01 | P<0.01 |
| Test 8 Delayed<br>Recognition | NS | P<0.01 | P<0.01 | P<0.01 | P<0.01 | NS |

The comparison of means scores between age groups is summarized in table 7. Whereas between adjacent age bins, there was no significant difference in individual test scores (at both extremes of age groups), significant differences were observed when the age groups were sufficiently apart, typically between the youngest two bins and the older bins.

**Table 7:** Test scores of each age group compared with older and younger groups using Mann Whitney U test

| Parameters | 40-49<br>vs<br>50-59 | 40-49 vs<br>60-69 | 40-49 vs<br>70-79 | 40-49<br>vs 80+ | 50-59<br>vs<br>60-69 | 50-59<br>vs 70-79 | 50-59<br>vs 80+ | 60-69<br>vs 70-79 | 60-69<br>vs 80+ | 70-79<br>vs<br>80+ |
| --- | --- | --- | --- | --- | --- | --- | --- | --- | --- | --- |
| Test 1 Verbal<br>Fluency | NS | P<0.01 | P<0.01 | P<0.01 | P<0.01 | P<0.01 | P<0.01 | NS | NS | NS |
| Test 2 Object<br>Naming | NS | P<0.01 | P<0.01 | NS | P<0.01 | P<0.01 | NS | NS | NS | NS |
| Test 3 Mental<br>State Examination | NS | P<0.01 | P<0.01 | P<0.01 | P<0.01 | P<0.01 | P<0.01 | P<0.05 | P<0.05 | NS |
| Test 4 Calculation | NS | P<0.01 | P<0.01 | P<0.01 | P<0.01 | P<0.01 | P<0.01 | NS | NS | NS |
| Test 5 Immediate<br>Recall | NS | P<0.01 | P<0.01 | P<0.01 | P<0.01 | P<0.01 | P<0.01 | NS | P<0.05 | NS |
| Test 6 Visuo-constructional<br>Ability | NS | P<0.01 | P<0.01 | P<0.01 | P<0.01 | P<0.01 | P<0.01 | P<0.01 | NS | NS |
| Test 7 Delayed<br>Word List | NS | P<0.01 | P<0.01 | P<0.05 | P<0.01 | P<0.01 | NS | NS | NS | NS |
| Test 8 Delayed<br>Recognition | NS | P<0.01 | P<0.01 | P<0.01 | P<0.01 | P<0.01 | P<0.01 | P=0.05 | NS | NS |

Table 8 summarizes the comparison between the four educational classes at each age level. We observed that scores differed significantly between educational groups in the younger age levels across all tests, but with increasing age, differences ceased to be significant in the object naming and delayed word list domains.

**Table 8:** Comparison between the four educational classes at each age bin, using Kruskal-Wallis Test

| Parameters | Within 40-49 | Within 50-59 | Within 60-69 | Within 70-79 | Within 80+ |
| --- | --- | --- | --- | --- | --- |
| Test 1 Verbal Fluency | P<0.01 | P<0.01 | P<0.01 | P<0.01 | P<0.01 |
| Test 2 Object Naming | P<0.01 | P<0.01 | NS | NS | P<0.05 |
| Test 3 Mental State Examination | P<0.01 | P<0.01 | P<0.01 | P<0.01 | P<0.01 |
| Test 4 Calculation | P<0.01 | P<0.01 | P<0.01 | NS | P<0.01 |
| Test 5 Immediate Recall | P<0.01 | P<0.01 | P<0.01 | P<0.01 | P<0.05 |
| Test 6 Visuo-constructional Ability | P<0.01 | P<0.01 | P<0.01 | P<0.01 | NS |
| Test 7 Delayed Word List | P<0.01 | P<0.01 | P<0.01 | NS | NS |
| Test 8 Delayed Recognition | P<0.01 | P<0.01 | P<0.01 | P<0.01 | P<0.05 |

## Discussion

The ethnic and educational diversity of the Indian population makes it necessary to obtain normative cognitive data for different regions. The most notable among the very few validated cognitive screening tools are those developed by Ganguli et al^3^ for rural Hindi speaking communities, and the Kolkata Cognitive Battery by Das et al.^5^ We now attempt to modify and tailor the KCB for Maharashtrian populations.

The GDS is designed such that high scores correlate with severe depression. Thus, the 90th percentile score of the GDS is used as the cut off to screen in individuals who are potentially clinically depressed. For our study sample, this cut off is 17 which is lower than the accepted standard score of 22^7^ in rural communities (the cut off in Das et al’s sample^5^ was 21). Our camp-based methodology was dependent on respondents being sufficiently ambulatory and motivated to come to the screening site, which may have excluded the most depressed individuals with the uppermost range of GDS scores. However, even this lower cut off showed a significant association with low cognitive scores. While the depressed tenth of the study group had statistically significant lower scores in each of the tests, the gap in mean scores was notably wide in the fields of immediate recall and verbal fluency. Research indicates that depression affects cognitive performance in older as well as younger subjects, affecting ability to complete complex tasks.^11^ Sheline et al,^12^ studying the effect of late life depression on cognitive performance, assessed five major cognitive domains in patients with major depression, and reported that changes in processing speed were found to most fully mediate the influence of predictor variables on all other cognitive domains.

Our study showed significantly poorer performance by women in each of the tests, compared to men. While the largest gender attributable score difference was observed in testing calculation, the difference in visuo-constructional ability scores was striking even in the lowest tenth percentile. The women were significantly less educated than men. Few among them (208/1316; 15.8%) had jobs outside the home, the majority being homemakers. The proportion of illiteracy was more than twice as high among women compared to men (f: 244/1316, 18.5%; m: 105/1335, 7.9%). The general awareness, and comfort with writing and drawing among women may have been adversely affected, as a result. Education, in particular literacy, has been shown to be associated with high cognition in the elderly.^13^ In a study of ethnically diverse elders, respondents with lower literacy levels had lower cognitive scores, this difference being attributed to poorer organization of visuospatial information, lack of previous exposure to stimuli, and difficulties with interpretation of the logical functions of language.^14^ Thus, it may be surmised that the observed differences in cognitive scores between men and women in our study are attributable to differences in their education levels.

Our study showed that test scores decreased (albeit not uniformly) with increasing age, with differences more pronounced between the youngest and oldest age groups; no significant difference being observed comparing the scores of the moderately older or younger age groups with each other. Das et al, in their Kolkata study, observed a similar pattern of significant differences in overall test scores mainly when comparing the relatively younger age groups (50-59) with elderly subjects aged above 70 years.^5^

The most remarkable finding of our study is the difference in test scores between the rural and urban subsets of the study group, particularly in the domains of visuo-constructional ability and immediate recall. Compared to rural, our urban respondents were significantly younger and better educated, with 50% of the urban dwellers having secondary level or higher education versus only 21% rural dwellers. The rural subset also had proportionately more women compared to urban (R: 791/1435; 55.1% vs. U: 525/1216; 43.2%). We noted a positive correlation of test scores with urban residence even after controlling for age, sex and education, with the correlation coefficients being highest in the domains of visuo-constructional ability (0.323, p< 0.01), immediate recall (0.313, p< 0.01), and calculation (0.239, p< 0.01). The modern, fast-paced urban life, involving daily commutes, monetary calculations, workplace interactions and a variety of cultural exchanges with different occupations and ethnicities, could have contributed to better general awareness, alertness and mental processing speed, and subsequently higher test scores. Researchers from other countries have noted a similar rural-urban gap in cognitive scores, attributing it to educational disadvantage, selection through migration, and the existence of potentially modifiable risk factors other than lifestyle.^15,16^ There is thus considerable scope for further, larger studies of cognitive differences in urban and rural India, exploring factors such as education, income, childhood illnesses and substance use habits.

### Ceiling effect

Das et al,^5^ in their study of cognitive scores in an urban elderly population, noticed possible ceiling effects in tests of object naming, calculation, mental state examination and delayed recognition, which they attributed to the large number of literate subjects in an urban milieu. However, no such effect has been observed in the present study, either overall or in the urban subset of the participants group. This may be consequent to the difference in the educational distribution of the two study sets, with two-thirds (501 of 745, 67%) of the Kolkata sample being educated at the secondary or higher level, compared to only a third of the present study group (898 of 2651, 34%).

### Limitations

A limitation of our study which may have introduced a bias, (in addition to the camp-based methodology which may have excluded the severely depressed) is that many of our older subjects were unable to recall their exact age, and there may have been marginal misclassification of age bins.

## Conclusion

We have presented and discussed the formulation and validation of a Marathi translation of the Kolkata Cognitive Battery. We found that cognitive scores declined with increasing age and poor education; however, our chief finding is the striking positive difference that an urban residential environment made to cognitive scores, when other variables were comparable. This Marathi KCB having been validated on a large sample of urban as well as rural residents, may be used to study cognition in Marathi speaking populations with appropriate cut-offs tailored to the degree of urbanization of the locality.

Our study participants performed poorly in the domains of verbal fluency, object naming and delayed recognition in comparison to the Ballabhgarh and Kolkata study groups with whom the original Hindi and Bengali cognitive testing instrument versions were validated. Further research is warranted to explore the factors leading to these differences. Women had poorer cognitive scores than men, which we surmise is due to their homebound lives, lesser education and lower involvement in financial management.

## Supporting information

STROBE checklist

Ethical approval document

## Data Availability

All data produced in the present study are available upon reasonable request to the authors

## Data availability statement

The data produced in the present study, and the Kolkata Cognitive Battery translated to Marathi (Marathi KCB) are available upon reasonable request to the authors.

