## Supplementary figures and images for "Normative cognitive scores in western India, stratified by age, rurality, cognitive domains, and psychiatric comorbidity"

### Ethical approval document

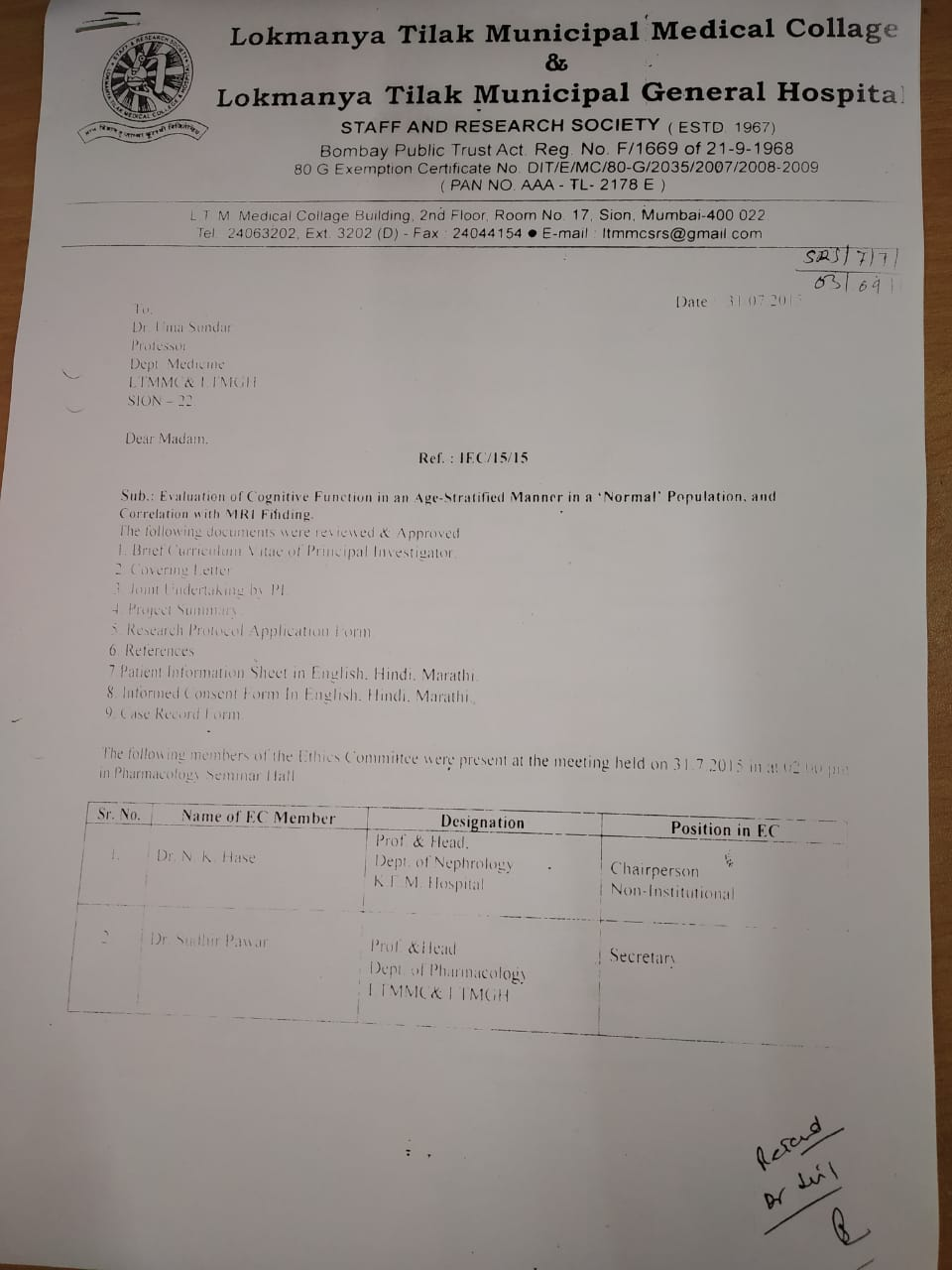

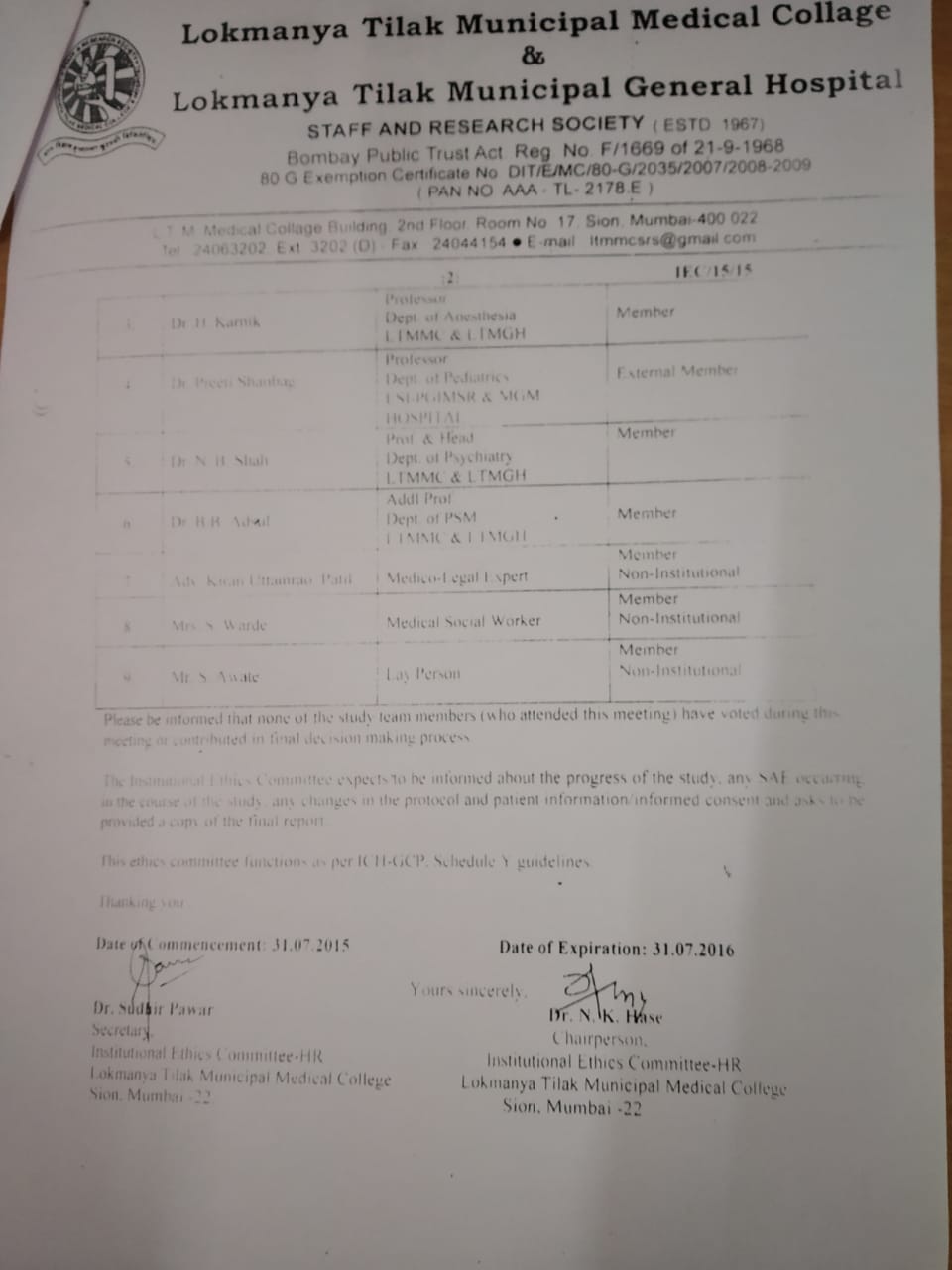
